# Using a Computational Cognitive Model to Simulate the Effects of Personal and Social Network Experiences on Seasonal Influenza Vaccination Decisions

**DOI:** 10.1101/2022.02.02.22269091

**Authors:** Matthew M. Walsh, Andrew M. Parker, Raffaele Vardavas, Sarah A. Nowak, David P. Kennedy, Courtney A. Gidengil

**Affiliations:** RAND Corporation, Pittsburgh, PA, USA; RAND Corporation, Santa Monica, CA, USA; RAND Corporation, Boston, MA, USA; Children’s Hospital Boston, Harvard Medical School, Boston, MA, USA

**Author notes:** Send correspondence to Matthew Walsh.

**Keywords:** Seasonal influenza, vaccination, cognitive model, public health, ACT-R

## Abstract

The substantial societal costs of seasonal influenza include illness, loss of lives, and loss of work productivity. Vaccination is the most effective means for averting the disease, yet fewer than half of adults in the United States are vaccinated annually. In this research, we focus on how personal experience and the experiences of close social contacts contribute to vaccination decisions. The results of a multi-year longitudinal survey study revealed the significant effects of personal and social network experiences on vaccination. We develop a memory-based model of vaccination decisions using the Adaptive Control of Thought – Rational (ACT-R) integrated cognitive architecture. The model accounts for the effects of personal and social experience on vaccination, and suggests interventions that may be effective in increasing vaccination uptake.

## Introduction

Seasonal influenza accounted for an average of 29 million cases, 440 thousand hospitalizations, and 36 thousand deaths in the U.S. each year from 2011 to 2020 (CDC, 2020a). The economic burden of influenza has been estimated at $11.2 billion dollars annually (Putri et al., 2018). Influenza vaccination is the most effective means of preventing the illness and averting its most severe outcomes (Thompson et al., 2018). Yet despite the relative efficacy and availability of the influenza vaccination, fewer than half of adults in United States are vaccinated each year (CDC, 2020b). Given the significance of this issue in terms of loss of lives and loss of productivity, it is essential to understand how people decide about vaccination in order to design interventions to increase vaccination uptake.

One of the strongest predictors of future health behavior, including vaccination, is past behavior (Ouellette & Wood, 1998). Past vaccination decisions are highly predictive of perceived risk of illness, intention to vaccinate, and future vaccination decisions (Gidengil, Parker, & Zikmund-Fisher, 2012; Maurer, Harris, Parker & Lurie, 2009; Walsh et al., 2020). Other factors like knowledge about influenza and vaccination, provider recommendation, personal experiences, social influences, and other sociocognitive variables are predictive of vaccination decisions as well (Brewer et al., 2017; Lehmann et al., 2015; Parker et al., 2013).

Research on health decision making has given rise to psychological theories such as the health-belief model, the theory of planned behavior, the theory of reasoned action, and the protection-motivation theory (Fishbein & Ajzen, 2010; Janz & Becker, 1984; Rogers, 1975). These theories propose that people appraise the probability and severity of health risks, the efficacy of recommended health behaviors, the barriers toward enacting those behaviors, and possibly the subjective norms surrounding them. Appraisals shape motivation, which in turn shapes behavior. Psychological theories of health behavior have been applied to vaccination decisions (for a review, see Brewer et al., 2017). Because these theories are broad, they account for many of the factors shown to be related to vaccination. But because these theories are also fairly abstract, they do not address finer-grained, dynamical changes in individuals’ behavior.

Computational models of vaccination behavior have also been developed (for a review, see Wang et al., 2016). These models can be divided among three categories. First, phenomenological models describe observed behavior effects mathematically without positing underlying mechanisms (i.e., Capasso & Serio, 1978). Although these models may be inspired by psychological factors, the mathematical equations describe outcomes rather than the specific psychological processes that give rise to them. Second, game theory models treat vaccination as a strategic interaction in which individuals seek to anticipate how others will behave and to make decisions that maximize their own outcomes (Bauch & Earn, 2004). Although game theory provides a useful framing for social aspects of vaccination, it makes the strong assumptions that individuals are driven by selfish motives and that they are rational decision makers. Third, psychological models invoke psychological theory to formulate mechanisms underlying vaccination decisions. For example, these mechanisms include learning, subjective expected utility estimation, and estimating the likelihood of infrequent events (Vardavas, Breban, & Blower, 2007). Despite their psychological origins, these models vary in terms of psychological fidelity and they are only indirectly linked to other established computational models from the fields of experimental and cognitive psychology.

In parallel and independently, research in cognitive science has sought to develop computational cognitive models that simulate cognitive processes (Walsh & Lovett, 2016). Many of these models have been developed and validated using data gathered in carefully controlled laboratory experiments. Yet the cognitive processes they represent are general. This raises the possibility that computational cognitive models can be used to understand and improve human behavior in real-world contexts (Gray, 2007). Human behavior in real world contexts, in turn, can provide new tests of computational cognitive models. The implications of these models with respect to vaccination decision making have not yet been explored, and is the focus of this paper.

Adaptive Control of Thought – Rational (ACT-R) is an integrated cognitive architecture that represents the cumulation of psychological theories encompassing human perception, memory, learning, judgement and decision making, and motor control (Anderson, Bothell, Byrne, & Douglass, 2004). ACT-R has a long history in educational psychology. By simulating student learning processes, ACT-R can be used to assess mastery and to tailor materials delivered to students by intelligent tutoring systems (Anderson et al., 1995). More recently, ACT-R has been used to develop interventions to increase uptake of health behaviors. For example, Pirolli et al. (2018) related components of ACT-R to the theory of planned behavior (2018). They then embedded ACT-R in a collection of mobile health applications, and used ACT-R to facilitate goal setting, to improve the timing of health behavior reminders, and to engender habit formation.

## Overview of the Current Study

The goal of this paper is to use a computational cognitive model developed in ACT-R to understand how personal experiences and the experiences of others in one’s social network influence seasonal influenza vaccination decisions. We conducted a multi-year longitudinal study during which individuals self-reported whether or not they vaccinated, and whether or not they contracted influenza during each season. Individuals also reported the vaccination behaviors and illness outcomes of up to 15 of their close social contacts. We used data from this study to address four questions:

1. How do recent personal experiences influence vaccination decisions?
2. How do recent experiences of close social contacts influence vaccination decisions?
3. How do the influences of past personal and social network experiences on vaccination decisions change over time?
4. Can ACT-R account for the longitudinal effects of personal and social network experiences on vaccination decisions?

Next, we describe a memory-based model of vaccination decisions implemented in ACT-R. We then report the results of the longitudinal survey study. Finally, we evaluate the model using data from the study. If ACT-R provides a valid account of vaccination decision making, then it could be used to prospectively explore the effectiveness of different interventions, and to select the ones that are most promising.

### A Memory-Based Model of Vaccination Decisions

At the core of ACT-R is an activation-based theory of declarative memory that accounts for the acquisition and retention of factual knowledge and experiences (Anderson et al., 2004). Information stored in declarative memory can be used to make decisions. For example, instance-based learning theory (IBLT), which uses ACT-R’s theory of declarative memory, proposes that when faced with a decision, people recognize the situation based on its similarity to past instances stored in memory (Gonzalez et al., 2003). They then use information stored in those instances to make decisions. The memory-based model of vaccination decision making that we propose directly follows ACT-R’s theory of declarative memory, and it uses declarative memory to enable decision making in the manner described by IBLT.

In ACT-R, information is stored in declarative memory as instances (or *chunks*). For example, an instance can include information about personal experiences, such as whether or not one vaccinated in the previous season and whether or not one contracted influenza. Instances are added to memory during different seasons and accumulated over time. An instance can also include information about observed experiences, such as whether or not a partner, friend, or co-worker vaccinated in the previous season and whether or not they contracted influenza.^1^ Once again, these instances are added to memory during different seasons and for multiple individuals in one’s social network.

Instances may be retrieved from declarative memory to enable decision making. In the case of vaccination, an individual may attempt to retrieve instances concerning past vaccination behaviors and influenza outcomes to inform the current decision of whether or not to vaccinate. These instances can be divided among eight classes based on three factors: (1) Whether they reflect personal experiences (ego) or social network experiences (alter); (2) Whether they involve vaccinating or not; and (3) Whether they involve contracting influenza or not (Table 1). The behaviors and outcomes listed in Table 1 refer to those of the individual (ego) or of others in their social network (alter). Each instance summarizes the combined effects of the multitude of experiences making up that class. Additionally, each instance provides evidence for or against future vaccination. For example, if an individual did not vaccinate and did not contract influenza in the previous season, that experience would provide evidence in favor of vaccinating again. Alternatively, if an individual vaccinated and did contract influenza, that experience would provide evidence against vaccinating again. In this instance-based model of vaccination decision making, the individual retrieves an instance from memory and makes a decision based on the evidence that it contains.

**Table 1.** Instances contained in model

| Source | Behavior | Outcome | Evidence |
| --- | --- | --- | --- |
| Ego | Vaccinate | Influenza | Not vaccinate |
| Ego | Vaccinate | Not Influenza | Vaccinate |
| Ego | Not Vaccinate | Influenza | Vaccinate |
| Ego | Not Vaccinate | Not Influenza | Not vaccinate |
| Alter | Vaccinate | Influenza | Not vaccinate |
| Alter | Vaccinate | Not Influenza | Vaccinate |
| Alter | Not Vaccinate | Influenza | Vaccinate |
| Alter | Not Vaccinate | Not Influenza | Not vaccinate |

Memory retrieval in ACT-R is competitive. The accessibility of an instance from memory depends on its activation value relative to the activation of the other instances stored in memory (Anderson & Lebiere, 1998). The activation value of an instance depends on three components: base-level activation (*B*_*i*_), spreading activation (*W*_*i*_), and mismatch penalty (*MP*_*i*_).

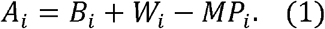

Base-level activation is a function of how frequently and recently an instance has been experienced,

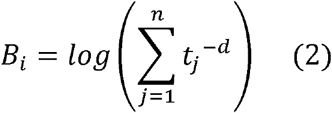

where *t*_*j*_ is the elapsed time since the instance was experienced on each of *j* occasions, and *d* is the decay rate. A particular instance, such as vaccinating and not contracting influenza, can be experienced multiple times (i.e., during each of multiple seasons, or for multiple alters during one season). The base-level activation of the instance reflects the summation across the collection of all such experiences.

Figure 1 illustrates the dynamics of base-level activation for three hypothetical scenarios. In the first scenario (top panel), an individual experiences the same outcome during Seasons 1, 2, 3, and 4. The activation of each experience decreases over time, but the cumulative activation increases with each repetition. In the second scenario (middle panel), an individual experiences the same outcomes during Seasons 2, 3, and 4. As compared to the first scenario, cumulative activation at the onset of Season 5 is somewhat lower due to the decreased frequency of the experiences. In the third scenario (bottom panel), the individual experiences the same outcomes during Seasons 1, 2, and 3. As compared to the second scenario, cumulative activation at the onset of Season 5 is somewhat lower due to the decreased recency of the experiences.

**Figure 1.**
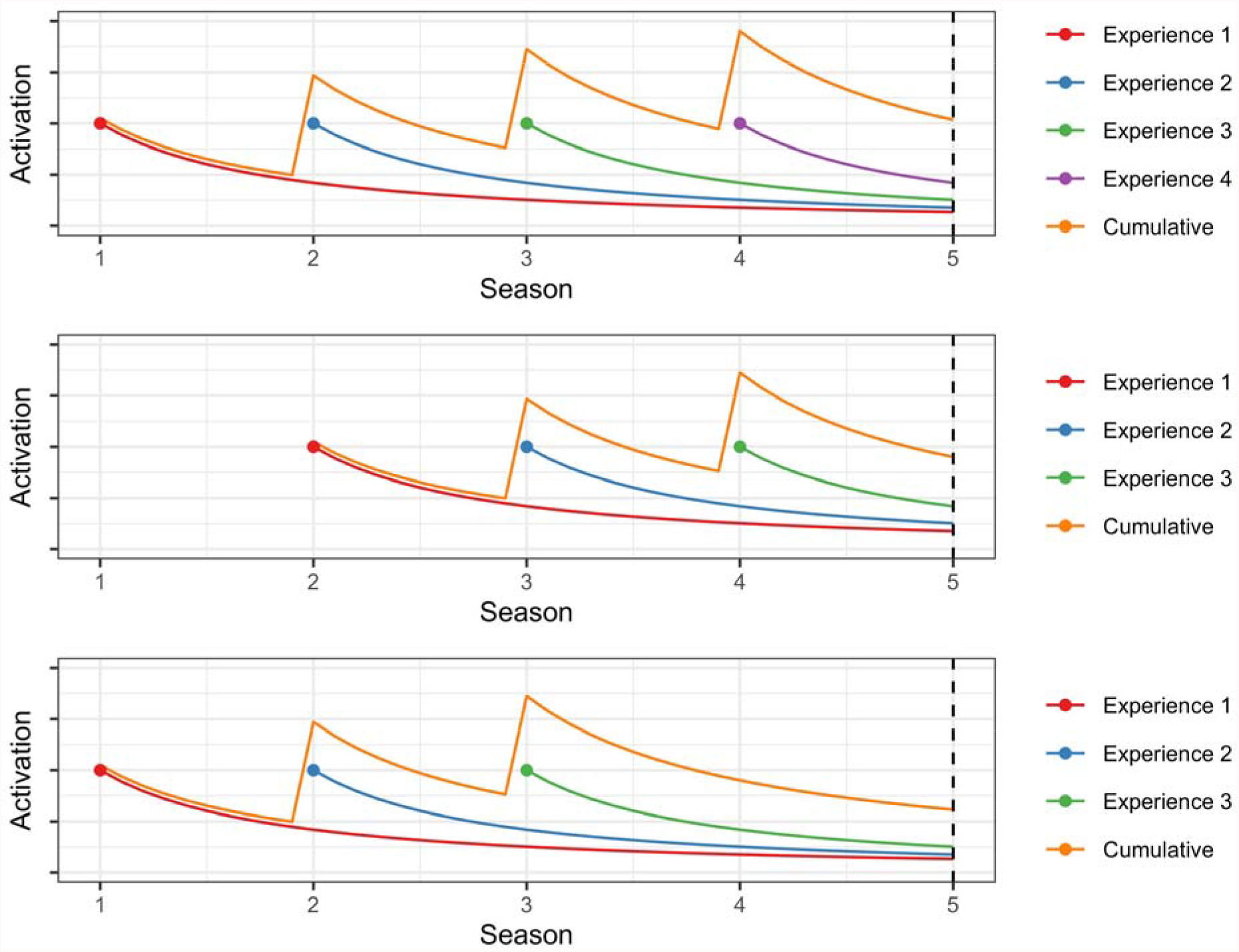
Activation as a function of frequency and recency of experiences

The accessibility of an instance from memory also depends on spreading activation, *W*_*i*_ (Eq. 1). Spreading activation describes how cues, present in the environment and associated with instances stored in memory, increase the accessibility of those instances from memory. Although cues can be directly manipulated in laboratory studies, they are unknown in the naturalistic context of our study. Consequently, rather than computing *W*_*i*_ from ACT-R’s spreading activation equation, we treated it as a free parameter that took separate values for the four types of instances formed by crossing vaccination decision (yes or no) and influenza outcome (yes or no).

Finally, the accessibility of an instance from memory depends on the mismatch penalty, *MP*_*i*_. An assumption in our model is that individuals seek to retrieve personal experiences (i.e., ego), but that they occasionally retrieve social network experiences instead (i.e., alter). This is how social network experiences exert an effect. However, because alters differ from the individual making the decision, the activation of alter instances (*A*_*i*_) are offset by the mismatch penalty. For example, if the mismatch penalties were set to zero for ego and alter instances, individual and social network experiences would be equally accessible from memory. However, if the mismatch penalty were set to zero and one for ego and alter instances, respectively, social network experiences would become somewhat less accessible from memory.

The activation values for instances are unbounded. The probability of retrieving an instance depends on its activation and is given by the equation,

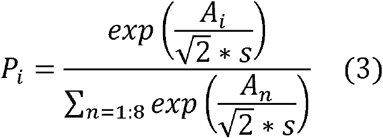

where *s* is the activation noise parameter. This equation converts unbounded activation values to retrieval probabilities for each of the eight instances.

In this instance-based model of vaccination decision making, retrieval probability is computed for each of the eight instances formed by crossing the three factors (Table 1). The probability of vaccinating equals the cumulative probability of retrieving one of the four instances that provides evidence in favor of vaccinating (and hence not one of the instances providing evidence against vaccinating). The model contains seven free parameters (decay, four unique spreading activation values, the mismatch penalty applied to alter instances, and activation noise), which are shared across individuals. These parameters are estimated from empirical data. The model predicts different vaccination probabilities for each individual based on their unique personal and social network histories.

## Methods

### Materials

From Fall 2016 to Spring 2020, eight online surveys were fielded longitudinally to a random sample from a panel of U.S. adults aged 18 years and older participating in the RAND American Life Panel (ALP), a nationally-representative panel of U.S. adults. The study protocols were approved by RAND’s Human Subjects Protection Committee. For more information about how ALP surveys are designed and fielded, see Pollard and Baird (2017).

The study used an egocentric social network interview protocol in which participants answered questions about themselves (the “ego”), they provided a list of network contacts (up to 15 “alters” such as family members, friends, and co-workers), and they answered questions about their perceptions of their network contacts’ characteristics and behaviors (Perry, Pescosolido, & Borgatti, 2018). At the start of the study, participants answered a question to generate a list of network contacts. Two surveys were then fielded during each influenza season, one in the fall and one in the spring. All spring surveys included a series of questions about whether the participant received the influenza vaccination during the previous season, as well as a question about whether they had an illness that they thought was influenza during the previous season. These questions were used to determine participants’ personal—or *ego*—experiences pertaining to vaccination and illness. Each spring survey also asked participants to update their list of social network contacts, and for each to report whether the person received the influenza vaccination and whether they had influenza during the previous season. These questions were used to determine participants’ social network—or *alter*—experiences pertaining to vaccination and illness. The survey questions used in the reported analyses are contained in the Supplementary Material.

Fall surveys, which are not analyzed in this paper, asked questions about participants’ expectations and attitudes toward vaccination and influenza. In addition, participants who had not completed the previous spring survey were asked about whether they had vaccinated and whether they had an illness that they thought was influenza during the previous season. Finally, the first survey included retrospective questions about the previous flu season, and so data from the surveys, which were administered over a four year period, encompassed a total of five seasons.

### Respondents

The first wave of the survey was administered to a total of 2,590 people, 2,173 of whom completed the survey (83.8% completion rate).^2^ All subsequent waves were administered to the sample of individuals who had completed the first wave (i.e., individuals who missed a given wave were invited to return in subsequent waves). Table 2 shows the number of participants contributing responses about personal experiences (ego) and about social network experiences (alter) by influenza season.^3^ Of the individuals who completed the social network survey, more than 85% reported perceptions for the maximum number (15) of social contacts permitted, and the rest reported perceptions for 1 to 14 social contacts.

**Table 2.** Sample size and self-reported vaccination and influenza rats for each surveyed influenza season

| Influenza Season | Sample Size |  |
| --- | --- | --- |
|  | Ego | Alter |
| Fall 2015 to Spring 2016 | 2,173 | 2,102 |
| Fall 2016 to Spring 2017 | 1,935 | 1,852 |
| Fall 2017 to Spring 2018 | 1,888 | 1,749 |
| Fall 2018 to Spring 2019 | 1,740 | 1,695 |
| Fall 2019 to Spring 2020 | 1,642 | 1,643 |

Of the participants, mean age in 2020 (the final year included in our analyses) was 60.1 years, 43% were male, 82% were white, and 47% had a bachelor’s degree or higher. A total of 1,486 provided complete personal and social network data from all five seasons, and 1,686 provided data from at least four of the five seasons. We retained participants with complete data from at least four seasons, and we used a K-Nearest neighbors approach to impute values from missing seasons.^4^

### Analyses

We begin by analyzing survey results to establish reported vaccination and influenza rates. We then examine the contributions of three sets of factors to individuals’ vaccination decisions: (1) Personal experiences from the previous season; (2) Social network experiences from the previous season; and (3) Personal and social network experiences from more distant seasons.^5^ Finally, we describe the computational implementation of the ACT-R model of vaccination and we apply the same analyses to the simulated data.

## Survey Results

### Reported Vaccination and Influenza Rates

The overall vaccination rate that participants reported for themselves was 54.4% and increased across survey seasons (Ego, Table 3), paralleling national increases in vaccination during that time (CDC, 2020b). Previous studies have revealed significant effects of demographics on vaccination behavior. To examine whether these effects replicated in our study, we analyzed individuals’ vaccination decisions during each season (yes or no) using a mixed effects logistic regression with gender, age, race (white versus non-white), education (bachelor’s degree or higher versus not) and survey year as fixed factors, and respondent as a random factor. Vaccination rate was higher in college educated individuals (*b* = 1.16, *z* = 4.69, *p* < .0001) and in white individuals (*b* = 1.27, *z* = 3.81, *p* < .0001), and it increased with age (*b* = 1.49, *z* = 10.940, *p* < .0001) and across survey year (*b* = 0.21, *z* = 7.37, *p* < .0001). Women vaccinated only slightly more frequently than men (*b* = 0.10, *z* = 0.42, *n*.*s*.).

**Table 3.**
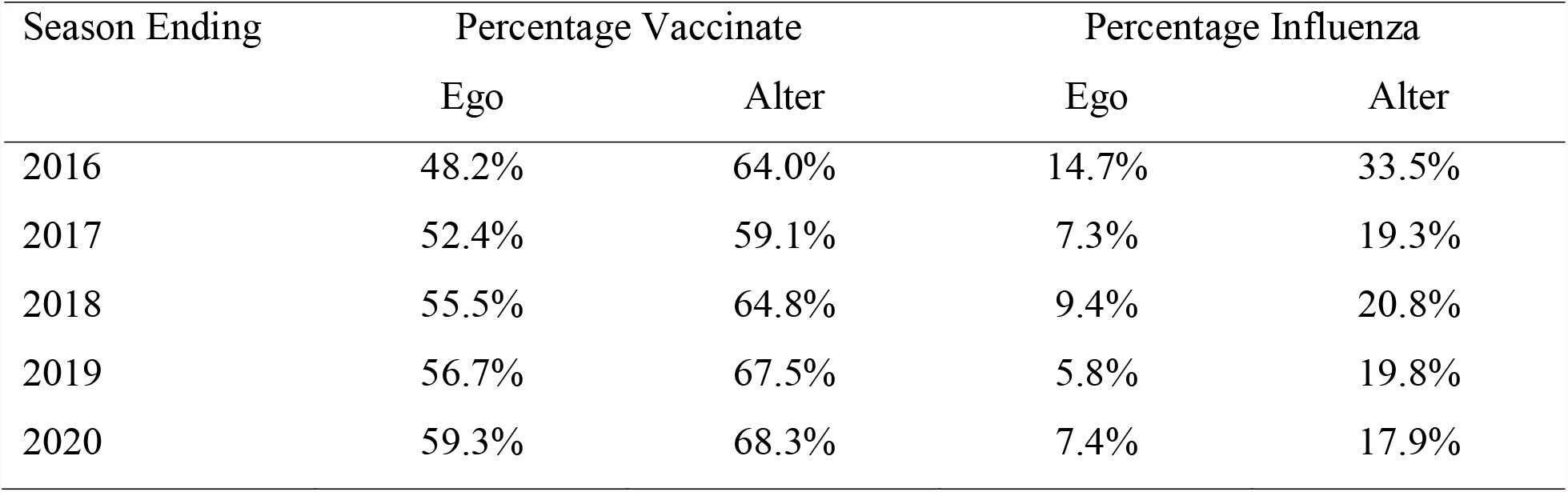
Vaccination rates and influenza rates reported for self (ego) and social network (alter) by influenza season

| Season Ending | Percentage Vaccinate |  | Percentage Influenza |  |
| --- | --- | --- | --- | --- |
|  | Ego | Alter | Ego | Alter |
| 2016 | 48.2% | 64.0% | 14.7% | 33.5% |
| 2017 | 52.4% | 59.1% | 7.3% | 19.3% |
| 2018 | 55.5% | 64.8% | 9.4% | 20.8% |
| 2019 | 56.7% | 67.5% | 5.8% | 19.8% |
| 2020 | 59.3% | 68.3% | 7.4% | 17.9% |

For each participant, we calculated the percentage of network contacts who they reported as vaccinating. The average of these vaccination percentages across participants was 64.7% and also tended to increase across survey seasons (Alter, Table 3).

The overall influenza rate that participants reported for themselves was 8.9% (Ego, Table 3). This decreased with age (*b* = -0.28, *z* = 4.80, *p* < .0001) and season (*b* = -0.22, *z* = 7.10, *p* < .0001), but did not vary by gender, ethnicity, or education. The average of the influenza percentages that participants reported for their social networks was 22.2% and also tended to decrease across survey seasons (Alter, Table 3).

### Vaccination Determinants

To determine the contributions of recent personal experiences to vaccination decisions, we computed the probability of vaccinating in the current season conditioned on an individual’s experience from the previous season. Figure 2 shows vaccination rates from the final season of the study. The figure also shows simulated vaccination rates, which we return to. Individuals overwhelmingly repeated the previous season’s behavior. However, contracting influenza increased the probability of vaccinating for individuals who had not vaccinated in the previous season by about 18.3 percentage points, whereas it decreased the probability of vaccinating for individuals who had vaccinated in the previous season by about 4.9 percentage points. We used mixed effects logistic regression to analyze vaccination decisions in the current season, treating the previous season’s vaccination decision (vaccinate or not), influenza outcome (flu or not), and their interaction as predictors along with participant’s age and gender. The analysis included data from all seasons in the study. Vaccination rates were higher for individuals who vaccinated during the previous season (*b* = 3.82 *z* = 48.87, *p* < .001), and for individuals who reported contracting the flu during the previous season (*b* = 0.40, *z* = 1.91, *p* < .1). The main effects were qualified by a significant interaction (*b* = -0.75, *z* = 2.88, *p* < .01). Contracting influenza increased propensity to vaccinate among individuals who previously did not, whereas it decreased propensity to vaccinate among those who previously did.

**Figure 2.**
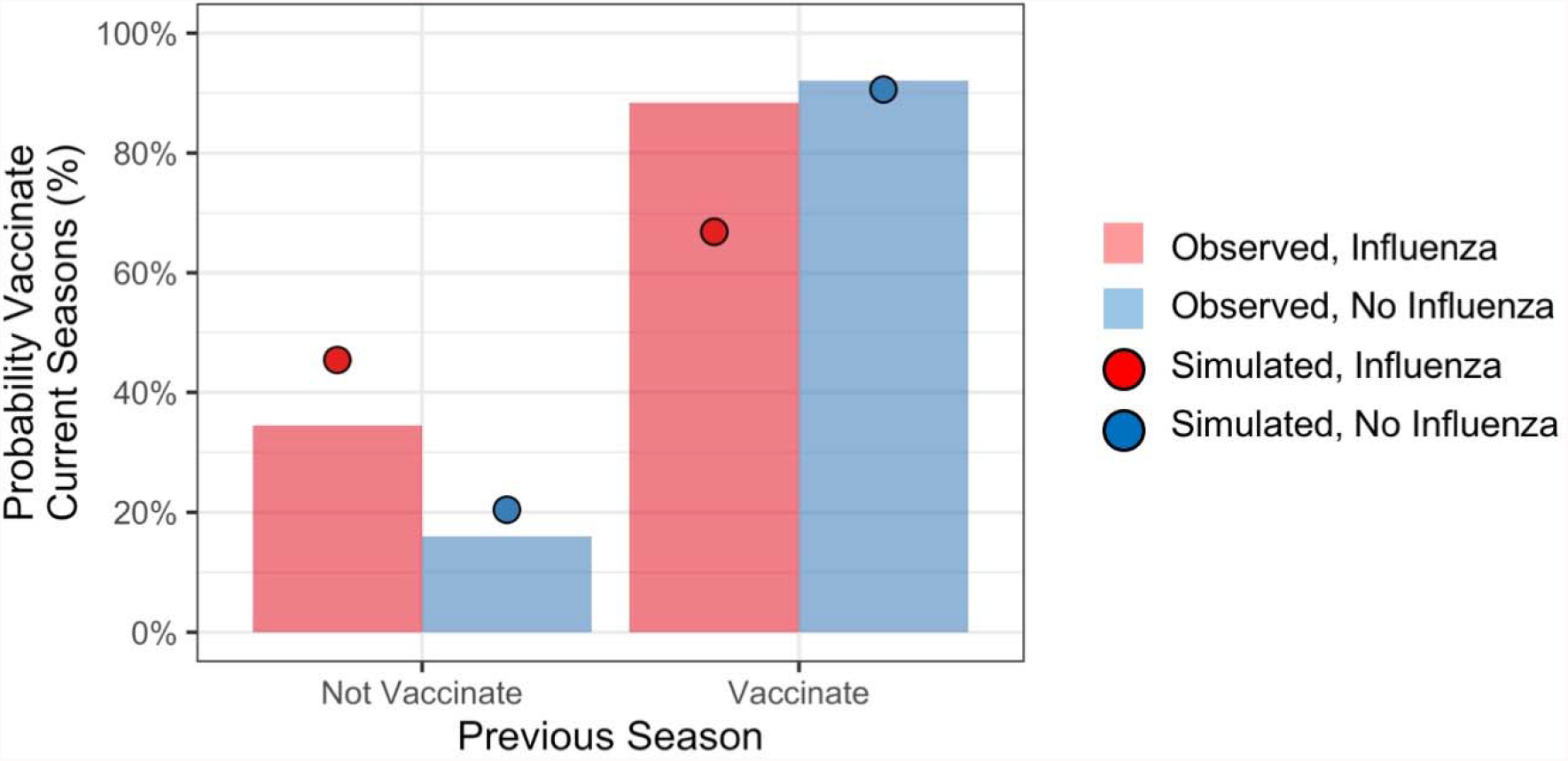
Probability of vaccinating in current season conditioned on personal experiences from the previous season

To determine the contributions of recent social network experiences to vaccination decisions, we computed the probability of vaccinating in the current season conditioned on the vaccination behaviors and influenza outcomes of alters from the previous season. Specifically, we computed the cumulative percentage of alters providing evidence that supported vaccinating (i.e., alters who vaccinated and *did not* contract influenza, along with alters who did not vaccinate and *did* contract influenza) and the cumulative percentage providing evidence that did not support vaccinating (alters who vaccinated and *did* contract influenza, along with alters who did not vaccinate and *did not* contract influenza). Figure 3 (red line) shows vaccination rates of participants from the final season of the study as a function of the percentage of their alters who provided evidence that supporting vaccinating during the previous season. Based on data from all seasons in the study, the probability of vaccinating increased with the strength of evidence from the social network component (*b* = 0.53, *z* = 3.74, *p* < .001).^6^

**Figure 3.**
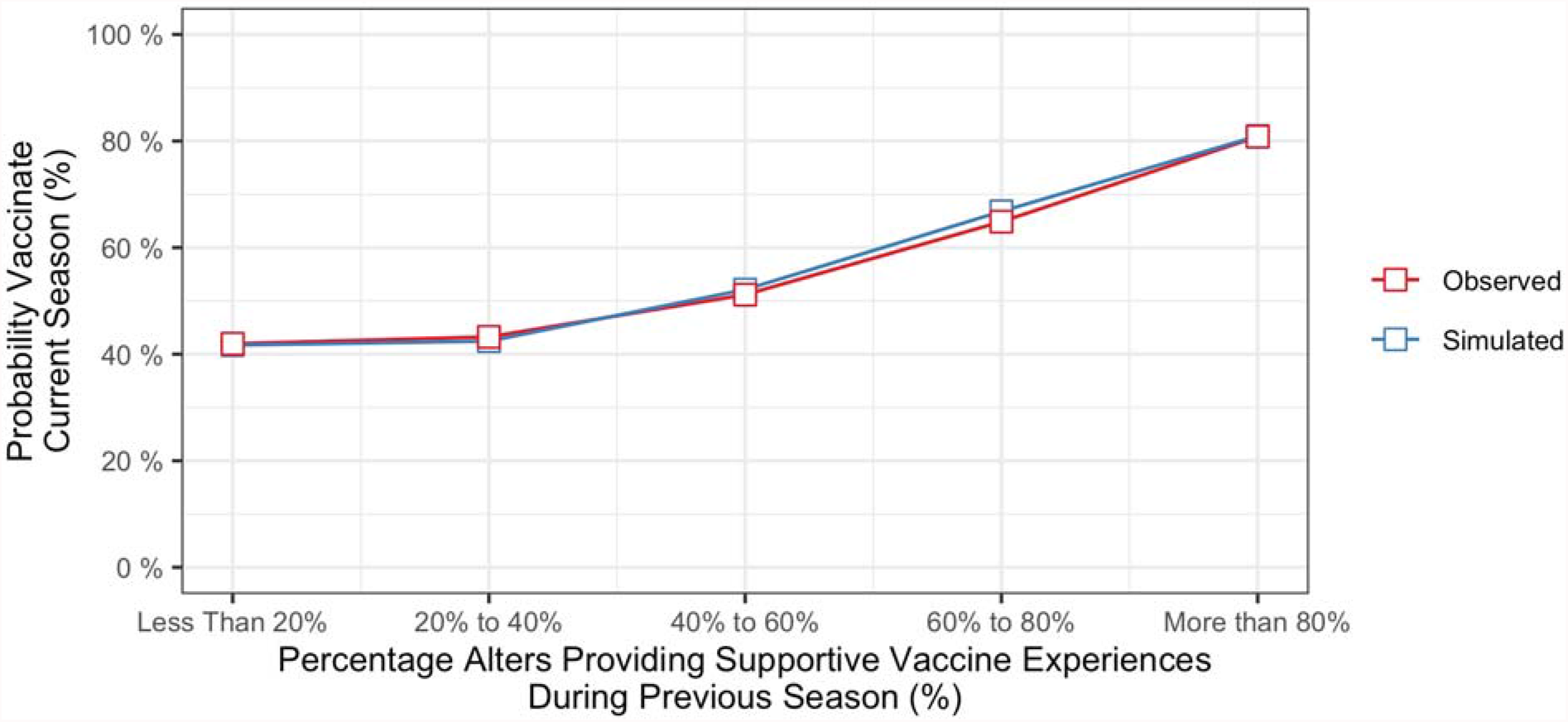
Probability of vaccinating in current season conditioned on evidence from social network from the previous season

Finally, to determine the contributions of more distant personal and social network experiences to vaccination decisions, we conducted a logistic regression treating individuals’ vaccination decisions in the final season of the study as the outcome, and their personal and social network experiences from each of the previous four seasons as the predictors. Figure 4 shows regression coefficients for personal and social network components as a function of elapsed time (years) since those experiences occurred (red curves).^7^ Coefficients were larger for personal experiences (red dotted line) than for social network experiences (red solid line), and coefficients decreased with elapsed time since those experiences occurred.

**Figure 4.**
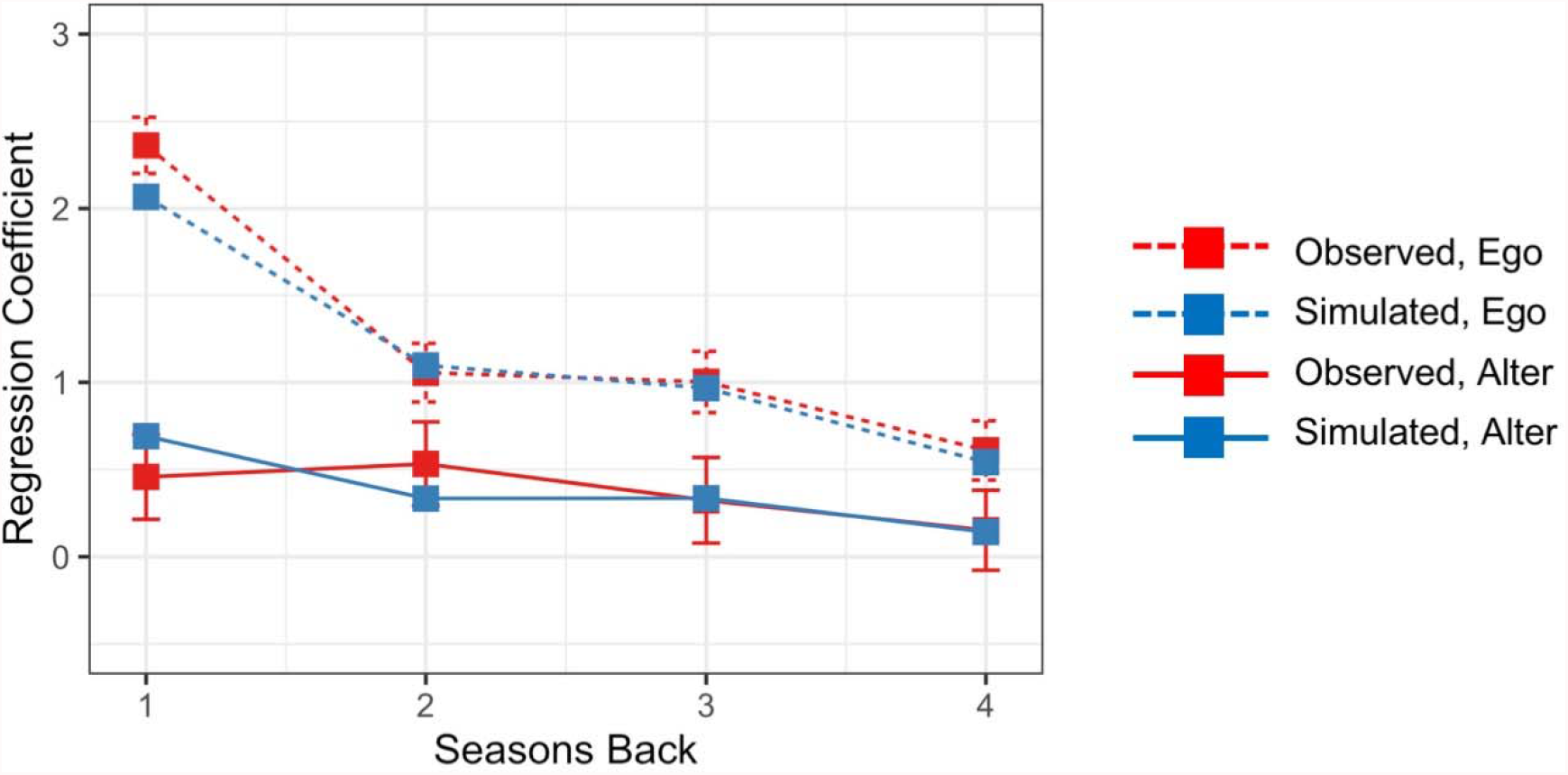
Regression coefficients for personal (ego) and social network (alter) experiences as a function of elapsed time (years) since those experiences occurred.

## Modeling and Simulation

### Model Fitting

We implemented the ACT-R memory-based model as a Bayesian hierarchical model and fitted it using fully Bayesian inference. Figure 5 expresses the model in graphical notation. We gave the model each respondent’s exact personal and social network experiences from the first four seasons of the study, corresponding to the period from Spring 2015 to Spring 2019. This is represented in the *alter* and *ego* nodes in Figure 5, which are shaded to denote that the values are observable. The dependent variable was the individual’s vaccination decision in the final season. This is represented by the *y* node in Figure 5, which is also shaded to denote that the values are observable. The model contains seven free parameters (*W*_*i*_, *d, MP*, and *s*).^8^ These appear as unshaded nodes in Figure 5 to denote that the values are not observable. A single set of values for the free parameters are estimated for and applied to data from all participants. The parameters are estimated to maximize the correspondence between the predicted probability of vaccinating and the observed outcomes given each participants’ history of personal and social network experiences. The model was implemented and fitted using the R interface to the Stan probabilistic programming language (Carpenter et al., 2017).

**Figure 5.**
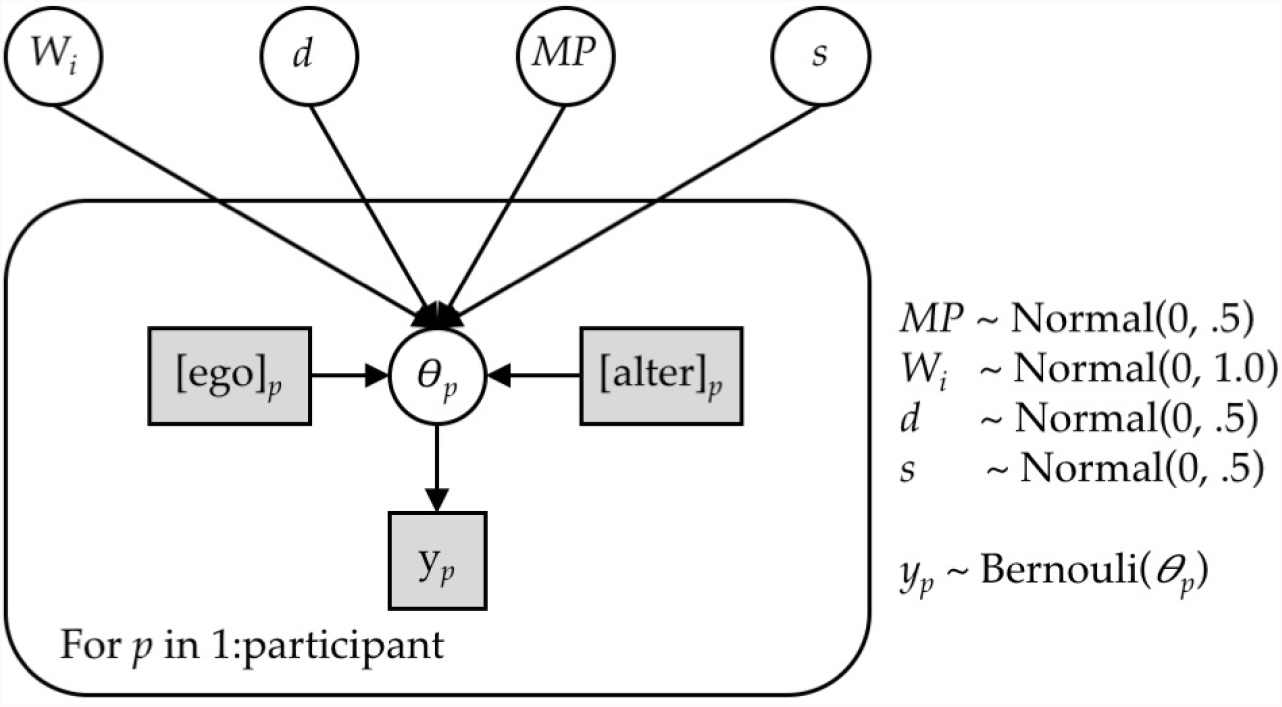
Graphical model for inferring vaccination decisions using instance-based model. Shaded boxes indicate the observable values.

Table 4 shows the means and standard deviations of parameter estimates. Given the high value of the mismatch penalty (*MP* = -3.21), an individual’s personal experience contributed far more to their behavior than the experience of any single alter. For example, the experience of personally vaccinating and not contracting influenza during the previous season produced an activation value that was slightly greater than 24 alters in one’s social network experiencing the same outcome.^9^ This means that individuals’ behavior was influenced far more by their own experiences than by the experiences of their social network. Yet given the probabilistic nature of memory retrievals, instances corresponding to alters could nonetheless be retrived. Further, given the non-linear relationship between the number of alters experiencing an outcome and the instance’s activation value, instances supported by a small number of alters still had a non-trivial probability of being retrieved.

**Table 4.** Parameter estimates

| Parameter | Name | Mean | SD |
| --- | --- | --- | --- |
| $MP$ | Mismatch penalty | -3.21 | 0.22 |
| $d$ | Decay | 1.16 | 0.19 |
| $s$ | Retrieval noise | 0.90 | 0.06 |
| $W_{vacc, infl}$ | Spreading activation: vaccinate/influenza | 0.21 | 0.18 |
| $W_{vacc, no infl}$ | Spreading activation: vaccinate/no influenza | 2.67 | 0.35 |
| $W_{no vacc, infl}$ | Spreading activation: no vaccinate/influenza | 0.40 | 0.30 |
| $W_{no vacc, no infl}$ | Spreading activation: no vaccinate/no influenza | 2.08 | 0.34 |

Given the moderate value of the decay parameter (*d* = 1.16), the strength of activation for instances that were one to two seasons old decreased from 1.00 to 0.45, and the strength of activation for instances that were three and four seasons old decreased further to 0.28 and 0.20. This means that the activation of an experience from the previous season was 2.2, 3.6 and 5.0 times greater than experiences from two, three and four seasons ago, respectively.

Finally, spreading activation varied by type of experience. Spreading activation was greater for experiences that did not involve contracting influenza (*W*_*vacc, no infl*_ = 2.67 and *W* _*no vacc, no infl*_ = 2.08). Surprisingly, spreading activation was near zero for experiences that did involve contracting influenza (*W* _*vacc, infl*_ = 0.21 and *W* _*no vacc, infl*_ = 0.40). Mechanistically, this means that outcomes that reinforced earlier behaviors (e.g., not contracting influenza) would be more retrievable from memory, and so would lead individuals to repeat those behaviors.

### Vaccination Determinants

We placed four seasons’ worth of historical data in the model’s memory and used it to generate vaccination probabilities for the final season. The mean of the model’s vaccination probability for the final season was similar to the observed vaccination rate (60.1% versus 59.3%). In terms of sensitivity, the mean of the model’s simulated vaccination probability for individuals who did vaccinate equaled 83.4%, and the mean for individuals who did not vaccinate equaled 25.1%.

The model produced different vaccination probabilities for individuals based on their personal experiences from the previous season (i.e., whether or not they had vaccinated, and whether or not they had contracted influenza). As shown in Figure 2 (red and blue circles), the model predicted that contracting influenza would increase propensity to vaccinate among individuals who previously did not, and it would decrease propensity to vaccinate among those who previously did.

The model also produced different vaccination probabilities for individuals based on their social network experiences from the previous season (i.e., whether or not an individual’s alters had vaccinated, and whether or not they had contracted influenza). As shown in Figure 3 (blue line), the model predicted that propensity to vaccinate would increase with the percentage of an individual’s alters providing vaccine supportive experiences during the previous season

Finally, the model produced diminishing effects of personal and social network experiences with the elapsed time since they had occurred. To evaluate this, we conducted a logistic regression treating the model’s predicted vaccination probabilities in the final season of the study as the outcome, and personal and social network experiences from each of the previous four seasons as the predictors. As shown in Figure 4, regression coefficients were larger for personal experiences (blue dotted line) than for social network experiences (blue solid line), and coefficients decreased with elapsed time since those experiences occurred.

To summarise, the ACT-R model corresponded reasonably well individuals’ vaccination decisions. In addition, it accounted for the effects of recent personnel experiences, recent social network experiences, and more distant personnel and social network experiences.

### Counterfactual Simulation

One surprising finding was the high values of spreading activation for experiences that involved not contracting influenza—that is, confirmatory outcomes (Table 4). As a result, personal and social network experiences involving not contracting influenza contributed disproportionately to decisions in the model. This raises the possibility that an intervention designed to increase retrievability of negative outcomes (i.e., contracting influenza) during seasons when an individual did not vaccinate may increase future vaccination uptake. To demonstrate the maximum potential of such an intervention, we conducted a counterfactual simulation in which spreading activation for personal and social network experiences that involved not vaccinating and contracting influenza was set to 2.08 (i.e., the same value as for not vaccinating and not contracting influenza). The simulated intervention increased the probability of vaccinating from 45.4% to 67.6% after seasons in which an individual did not vaccinate and they contracted influenza, (Table 5).

**Table 5.**
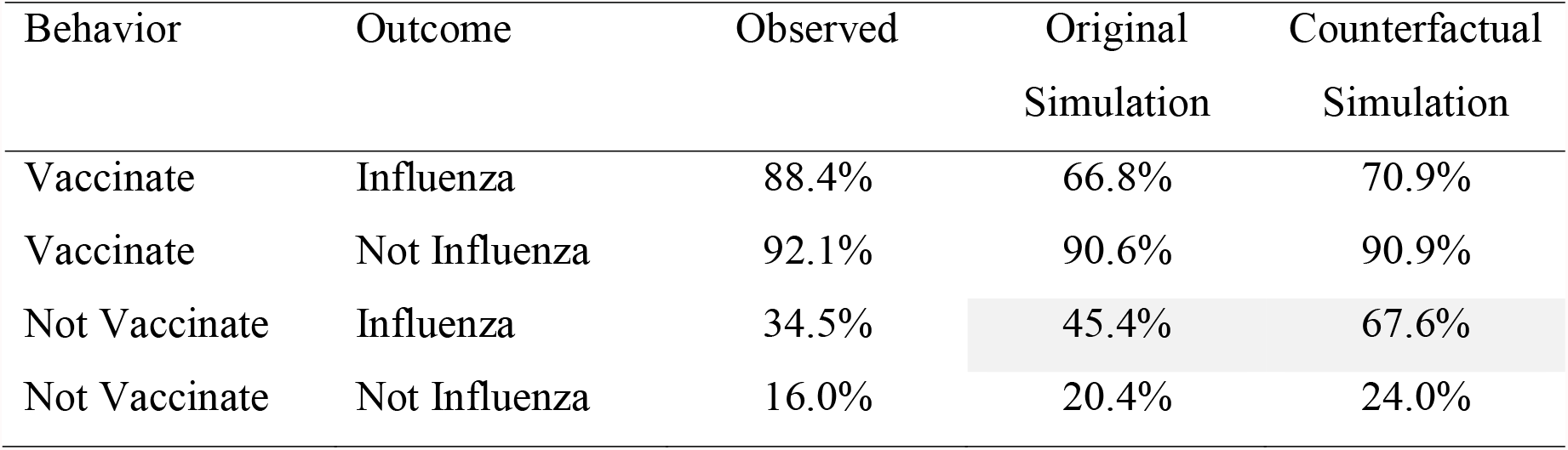
Observed and model vaccination rates

## Discussion

The goal of this study was to understand how personal experiences and the experiences of close social contacts contribute to vaccination decisions, and to test whether an ACT-R memory-based model could account for vaccination decisions. The results of this study support three conclusions, which we discuss in turn.

First, personal experiences strongly contributed to vaccination decisions. In line with earlier findings, people who previously vaccinated tended to do so again, and people who did not previously vaccinate tended not to in the future (Walsh et al., 2020). Survey responses and model estimates for the spreading activation parameters converged on the idea that propensity to repeat the previous behavior was strongest after experiencing the desired outcome (i.e., not contracting influenza). Yet there was some evidence that people learned from negative experiences as well (Jin & Koch, 2018; Walsh et al., 2020). After contracting influenza, individuals who had not vaccinated were slightly more likely to do so in the future, whereas individuals who had vaccinated were slightly less likely to do so again.

The finding that model-based estimates for spreading activation were so much greater for experiences that involved not contracting influenza versus ones that did was somewhat surprising. Practically, this is because individuals were only somewhat less likely to repeat their prior decision after contracting influenza (c.f. Figure 2). As shown in the counterfactual simulation, increasing spreading activation for one negative outcome (contracting influenza after not vaccinating) did produce a stronger behavioral reversal. This surprising finding is consistent with research showing that unpleasant personal experiences are recalled less well over time than pleasant ones (Walker & Skowronski, 2009). This finding is also consistent with research on the confirmation bias, which describes the tendency for people to seek evidence that confirms their prior beliefs (Nickerson, 1998). With respect to vaccination decisions, individuals searching for online health information select sources consistent with their prior beliefs and they rate belief-consistent information as being more credible and useful (Meppelinck et al., 2019). Our finding suggest that individuals may also recall personal experiences that are consistent with their prior vaccination beliefs, as revealed by their past vaccination decisions.

Second, the experiences of close social contacts contributed to vaccination decisions. Once again, individuals were sensitive to the combination of behaviors and outcomes rather than the simple base rates of vaccination and illness. As the percentage of social contacts providing supportive vaccine experiences increased, so too did the probability that the individual would vaccinate. Notwithstanding the statistical and practical significance of this effect, the regression analysis and the model estimate for the mismatch penalty converged on the idea that individuals were influenced more strongly by personal experiences than by social network experiences.

Earlier studies that examined the social nature of vaccination decisions have established the tendencies for people to choose to associate with similar individuals (homophily) and for trends to travel across social networks (contagion). Because of these processes, people tend to share characteristics with other close social contacts (Christakis & Fowler, 2013). Indeed, participants in our study who vaccinated also reported higher vaccination rates among close social contacts than participants who did not vaccinate. However, the base rate of vaccination among close social contacts alone did not strongly predict people’s decisions in our study. Rather, the health outcomes of close social contacts, conditional on their health behaviors, were far stronger predictors of vaccination decisions. This suggests that the social network effects arose from social contacts’ experiences and not just from their shared characteristics.

Third, the regression analysis and the model estimate for the decay parameter converged on the idea that the most recent personal and social network experiences exerted the strongest influence on decisions. However, more distant experiences continue to exert a weak but persistent effect. This helps to explain why individuals’ behavior do not change more dramatically after they experience a single negative outcome (i.e., contracting influenza). The recent experience is partially offset by the weight of past experiences.

Traditional theories of health behavior, though conceptually grounded, are too abstract to account for fine-grained, dynamically changes in individuals’ behavior. Computational models of vaccination behavior on the other hand are mathematically precise, but they are only indirectly linked to theories of psychological processes. Cognitive architectures like ACT-R offer alternative models based on a realistic characterization of human cognition.

As shown in this study, the outputs of the ACT-R memory-based model closely matched the empirical results. This supports its potential as an alternate to other theoretical and computational models of vaccination behavior. There are several benefits to using such a model. For example, ACT-R is a general theory of cognition that has been extensively validated in laboratory experiments and, increasingly, in real-world settings. Leveraging pre-existing components of ACT-R places vaccination within a set of general cognitive processes and bypasses the need to specify new mechanisms to account for vaccination behavior. Additionally, ACT-R offers the opportunity to situate a computational model of vaccination behavior alongside computational models of other health behaviors created using the same integrated cognitive architecture (Pirolli et al., 2018). Finally, the mechanistic nature of the ACT-R model allows it to account for existing data and to prospectively simulate outcomes of different interventions and policy options. As seen in our counterfactual simulation, an intervention that increases retrievability of negative outcomes that occur after failing to vaccinate may be effective. Other interventions suggested by the model, like reminding individuals about personal and social network experiences that support vaccinating, may increase vaccination rates as well.

Although the ACT-R memory-based model closely matched the empirical results, it is not a complete model. It does not represent certain factors known to influence vaccination; for example, provider recommendation and mass media (for a review, see Brewer et al., 2017). Additionally, the model treats all social contacts equally, whereas the influence of especially close social contacts (e.g., a spouse or family member) might be expected to be greater than the influence of a more distant ones (e.g., a co-worker or friend). This could be addressed by varying the mismatch penalty for different contacts depending on relationships. Finally, the model does not directly account for decisions made in the absence of personnel experience. This may be the case for illnesses that have been largely eradicated such as diphtheria and polio, which people nonetheless receive vacciantions for. This may also be the case for novel viruses. For example, when viruses like H1N1, Ebola, and COVID-19 emerge, new vaccines are developed and offered to the public. Because the viruses and vaccines are novel, individuals do not yet have a history of experiences with them. In these cases, individuals may generalize from related experiences (i.e., influenza vaccination), or they may rely on other sources of information to make decisions from descriptions rather than from experiences.

The results of this study must be considered in light of the fact that vaccination behavior and influenza outcomes were measured using self-report rather than actual medical records, and so may be susceptible to self-report biases. Self-reports of vaccination behavior have high agreement with medical records (Irving et al., 2009; Mangtani, Shah, & Roberts, 2007). However, self-reports of contracting influenza, and reports of vaccination behavior and influenza outcomes of close social contacts are likely less accurate. This may underlie the different vaccination and influenza rates that individuals reported for themselves versus for their social contacts (Table 3). Nonetheless, in terms of a memory-based model, it makes sense that if people’s perceptions are inaccurate, their decisions will be based on those inaccuracies rather than on ground truth.

Computational cognitive architectures are intended to represent general theories of cognition. The results of our study provide support for one computational cognitive architecture, ACT-R, in the novel context of vaccination decision making. In addition, these results demonstrate the potential for using ACT-R to understand vaccination decision making and to increase vaccination uptake.

## Data Availability

All data produced in the present study are available upon reasonable request to the authors

https://alpdata.rand.org/

## Supplementary Materials

### Survey Questions

The first wave of the survey was administered in Fall 2015 (Supplementary Table 1). The survey included two questions asking participants to retrospectively report the number of years since they last got the influenza vaccine and the number of years since they last had an illness that they thought was the flu. We coded these responses to reflect the participants’ vaccination and illness experiences from the previous season. The first wave of the survey also asked participants to identify up to 15 close social contacts. They were given the prompt, “From time to time, most people discuss important matters with other people. Looking back over the last several years, who are the people with whom you discussed matters important to you? Please list up to 15 of these people individually. Please only consider people who are 18 years old or older.” Finally, for each social contact identified, participants were asked to state how strongly they suspected that they had caught the flu and got the flu vaccine. Participants chose from five response levels ranging from “Definitely yes” to “Definitely no”. We coded responses of “Definitely yes” and “think yes” as *yes*, and we coded responses of “Definitely no” and “think no” as *no*.

The second wave of the survey was administered in the Spring 2016 (Supplementary Table 1). This survey and all later Spring surveys included a sequence of questions asking participants about whether they vaccinated in the previous season in response to a provider recommendation and, if not, whether they vaccinated without a recommendation. The survey also asked participants whether they had an illness they thought was influenza during the previous season. Finally, participants were asked how strongly they suspected that each of the social contacts they listed had caught influenza and got the influenza vaccination. These responses were coded in the same was as for the first wave of the survey.

To summarize, a total of eight surveys were administered over four years. Because the first survey included retrospective questions about the previous influenza season, data from the surveys covered a total of five years.

**Supplementary Table 1.**
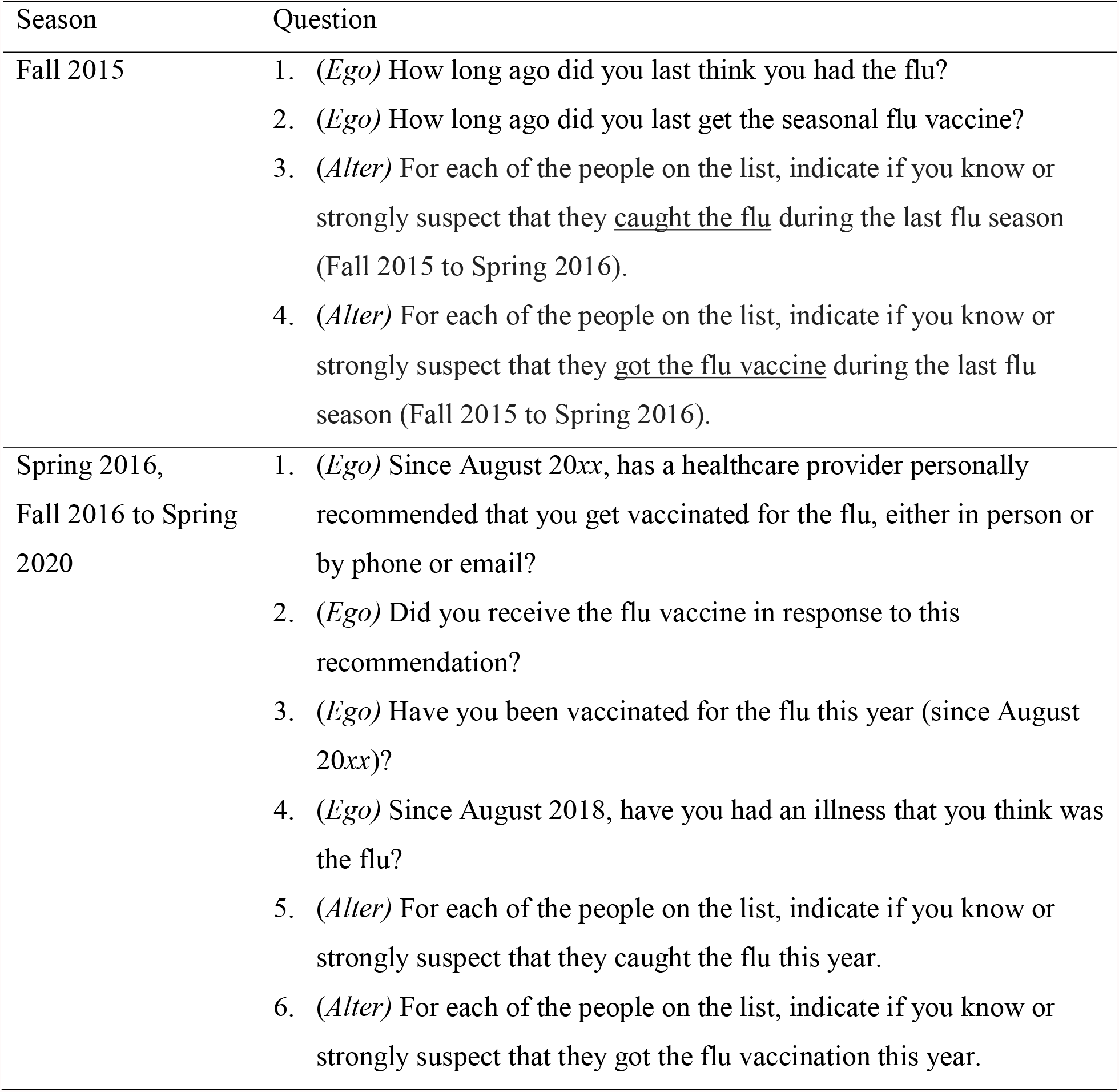
Vaccination and Influenza Questions Included on Survey

### Disclaimer

Conclusions drawn from this working paper do not necessarily represent the opinions of the RAND Corporation.

## Acknowledgements

This study was funded through support from the National Institute of Allergies and Infectious Diseases (R01AI118705). The views expressed are those of the authors and do not necessarily represent the views of these funders. In addition, data from previous surveys on the American Life Panel, funded by other sponsors, were incorporated to provide longitudinal vaccination estimates. For additional information see https://alpdata.rand.org/.

## Footnotes

1 Our model does not distinguish between different types of “observed” experiences, such as witnessing a social contact’s illness versus hearing about their illness.

2 Some individuals attrited during the first year of the study, and so all subsequent waves were administered to the 2,168 individuals who completed the first year of the study.

3 The number of ego responses is higher because participants were allowed to retrospectively complete those questions during the fall survey if they had missed the previous spring survey. Participants were not allowed to retrospectively complete social network questions.

4 We imputed missing values for whether or not a participant vaccinated, whether or not they had an illness that they thought was influenza, and whether or not their alters vaccinated and/or had influenza. The imputed values were based on their averages from the K other instances (i.e., participant-by-year records) most similar to the instance with missing values (*K* = 5). Similarity was based on demographic variables (age, gender, education, and race) along with ego and alter data from all completed surveys. We used the caret package in R to perform the imputation.

5 Different individuals reported different numbers of close social contacts. To give equal weight to each individual, we converted social network reports into the percentages of an individual’s close social contacts who vaccinated and who contracted influenza.

6 Each point in the figure reflects a mixture of individuals, some of whom vaccinated and some of whom did not. The individual differences that remain after controlling for social network experiences reflect, in part, differences in personal experiences.

7 Personal experiences were coded as supporting or not supporting vaccination (1 or 0), and social network experiences were coded as the percentage of alters providing vaccine supportive experiences.

8 The spreading activation parameter, *W*_*i*_, takes four values (*i* = 4) corresponding to the four outcomes formed by crossing vaccination decision (yes or no) and influenza outcome (yes or no).

9 For example, based on Equation 1, the activation of the instance resulting from personally vaccinating and not contracting influenza is 2.67. The activation of the instance resulting from 24 alters vaccinating and not contracting influenza is 2.64.

